# Educational attainment among primary school children with neurodisability: A population-based cohort study using linked education and health data from England

**DOI:** 10.1101/2025.06.12.25329491

**Authors:** Ayana Cant, Ania Zylbersztejn, Laura Gimeno, Vincent Nguyen, Joachim Tan, Ruth Gilbert, Katie Harron, the HOPE Study Group

## Abstract

**Objective:** To support integrated health and education approaches by assessing differences in educational attainment between primary school children with and without a range of neurodisabilities.

**Design:** Population-based cohort study.

**Methods:** We derived a national birth cohort of 2,351,589 children born in England between 01/09/2003-31/08/2008 enrolled in state-funded primary schools in Reception (age 4-5) using linked health and education records. Neurodisability was identified from hospital admission records. We described differences in primary school educational attainment for children with and without neurodisability.

**Results:** 2.2% of children had a recorded neurodisability before starting Reception. These children consistently underperformed in national assessments, with fewer than half meeting nationally expected levels in Maths and English at every time point. By the end of primary school (age 10/11), 31% of children with neurodisability did not participate in national assessments despite being enrolled in school. Among children with neurodisability, educational attainment was lowest for children with Down syndrome and highest for children with perinatal conditions.

**Implications:** Substantial attainment gaps exist between children with and without neurodisability. By the end of primary school, many children with neurodisability are excluded from being formally assessed, highlighting the need for more personalised, functional outcome measures to ensure the meaningful evaluation of their educational development.

**KEY MESSAGES:** What is already known on this topic

Children with neurodisability face functional challenges that can disrupt their success in school.

There is limited understanding of their participation in national assessments and academic attainment throughout primary school on a population level in England.

What this study adds:

Large attainment gaps exist between children with neurodisability and their peers. Most (70% of) children with neurodisability who enter Year 1 are not ‘school ready’. A third of children with neurodisability are excluded from assessments at the end of primary school despite being enrolled in school.

How this study might affect research, practice or policy:

Health records can identify children with neurodisability early, supporting timely special educational needs and disability (SEND) assessments before school starts. Joint support provided by health and education systems may reduce the burden of neurodisability on school outcomes.

## INTRODUCTION

Education aims to promote the socioemotional, cognitive and physical potential of every child. However, chronic health conditions in childhood can disrupt educational experiences, placing affected children at risk of poorer outcomes in later life.^1^ As a social determinant of important lifelong outcomes, education is a key factor to be prioritised for children with chronic conditions to promote their health, wellbeing and societal participation.

Neurodisability refers to a set of chronic conditions attributed to impairments of the nervous system, leading to functional limitations in movement, cognition, hearing, vision, communication, emotion, or behaviour.^2^ Children with neurodisability often face complex healthcare and educational needs. Neurodisability includes neurodevelopmental disorders (e.g., attention-deficit/hyperactivity disorder [ADHD], autism spectrum disorder [ASD], learning disabilities), neurological disorders (e.g., epilepsy, cerebral palsy), as well as a broader range of genetic, muscular, and perinatal conditions that affect brain development and learning.^1^ While individual conditions are rare, neurodisability affects around 1 in 25 (3.6%) primary school children born in England when considered collectively.^3^

Population cohort studies have been conducted in various settings (e.g. Scotland,^4,5^ Wales,^6^ Australia^7^) and indicate that neurodisability is associated with lower academic performance, higher rates of school absenteeism, and an increased need for additional learning support than unaffected peers.^4–8^ The disparity in educational attainment has been linked to multiple barriers to full engagement in education, including absences for health-related reasons, physical barriers to classroom access, communication difficulties, cognitive impairments, and challenges with emotional and behavioural regulation.^9–11^ Existing research often focuses on specific neurodisability conditions in isolation, rather than examining the broader experience of affected children across the school-aged population. There is a lack of population-level evidence for this group of children and their educational attainment in England.

Quantifying differences in attainment between children with neurodisability and their peers can provide prognostic information about expected educational attainment for those able to participate in the standard educational curriculum and those who are not formally assessed. Such insights can be used to plan local and national education strategies to meet the needs of children with neurodisability in mainstream or special education and inform joint working between health and education providers.

In this study, we used the Education and Child Health Insights from Linked Data (ECHILD) database of linked education and hospital records for all children in England^12^ to describe educational attainment throughout primary school at ages 5, 7 and 11 for children with neurodisability compared to their peers, and for five subgroups of neurodisability (neurodevelopmental disorders, epilepsy, cerebral palsy, Down syndrome and perinatal conditions).

## METHODS

A protocol for this study was published in March 2024 (Supplementary Materials A).^13^

### Participants

We used the ECHILD database which primarily links administrative data from the National Pupil Database (NPD) and Hospital Episode Statistics (HES).^12^ The version of ECHILD that we used for this study contained data on approximately 14.7 million children and young people aged 0-24 in England who were born between 1st September 1995 and 31st August 2020.^13^ Education data from the NPD contains information on children attending state-funded schools in England for academic terms from 2001/2 onwards.^14^ This includes pupil-level information on registration, attainment, exclusions, absences of all children attending state-funded schools in England. HES records all hospital admissions in National Health Service (NHS) funded hospitals in England (including birth), capturing 98-99% of all hospitalisations.^15^ HES was deterministically linked by NHS England to the NPD.^14^

The study included all singleton children born in NHS-funded hospitals between 1^st^ September 2003 and 31^st^ August 2008 linked to the NPD (Figure S1). Children were included if they appeared in the January (Spring) School Census of Reception (primary school entry; age 4/5) at a state-funded school. Children were followed from primary school entry in Reception until the end of Year 6 (age 10/11) or death, whichever occurred first (Figure S2). Cohort derivation details are included in the Supplementary Material B.

The research project was informed by children, young people, parent-carers and professional groups to understand their experiences of school and views on the use of linked health, education, and social care data for research. In focus groups and interviews, participants emphasised the importance of including the whole population to investigate the interrelated areas of health and education.^16^ We learned that exams and performance under the National Curriculum at school were points of stress for children with health conditions, forming a key outcome measure in this study. Key learnings from past public engagements can be found at: https://www.echild.ac.uk/engaging-with-the-public

### Hospital-recorded neurodisability

Neurodisability, as identified in hospital records, was defined by the presence of relevant diagnostic or procedure codes in hospital admissions from birth to the start of Reception, or as any cause of death in mortality records. We included conditions likely to involve neurodisability, where ≥50% of affected children would be expected to experience functional limitations and neurological impairment. The development and validation of the neurodisability code list have been described in detail elsewhere and the full list of codes and conditions is made available via the ECHILD phenotyping library.^3,17^ Children without records of neurodisability were classified as unaffected peers.

### Outcome: Primary school attainment

We evaluated primary school attainment at the end of Reception, termed the Early Years Foundation Stage Profile (EYFSP, age 4/5), at Key Stage 1 in Year 2 (KS1, age 6/7) and at Key Stage 2 in Year 6 (KS2, age 10/11) in nationally administered, universal assessments. At each Key Stage, we derived a) the proportion of children who did not complete the assessment to be taken in that year (i.e. appear in the Census but did not have an assessment record) and of those who did; b) the proportion of children, out of all those in the Census in that year, who reached nationally expected levels in these assessments as defined in line with Department of Education standards; and c) the cohort-specific standardised test scores calculated using the mean and standard deviation of all children’s scores who were assessed in a given academic year. We derived Maths and English results from the NPD at all three timepoints, as well as the ‘Good Level of Development’ (GLD) measure which is specific to the EYFSP. The GLD indicates whether a child is on track in key areas of learning before they enter Year 1 (age 5/6), including physical development, socioemotional development, and communication.^18^ Details on the derivation of primary school attainment outcomes are provided in Table S1.

### Statistical analysis

We described the derivation of the study cohort, the prevalence of neurodisability by entry into Reception and the proportion of children with and without neurodisability according to demographic and socioeconomic characteristics. This included sex assigned at birth, neighbourhood deprivation (quintile of Income Deprivation Affecting Children Index; IDACI), recorded entitlement to free school meals (FSM), ethnic group, month of birth, and region of residence. We also considered maternal age at delivery, gestational age at birth, and birthweight. A full description of the covariates is provided in Table S2.

For children with and without neurodisability, we described the proportion of children in the school Census in Reception, Year 2, or Year 6 who were: a) not assessed under the National Curriculum; b) were assessed but did not reach nationally expected levels; and c) were assessed and did reach nationally expected levels. We also calculated the mean (SD) standardised test scores for Maths and English out of all children who were assessed in each year.

We examined relative differences in educational attainment by comparing the likelihood of not meeting nationally expected levels (including both non-assessment and being assessed but not achieving) between children with neurodisabilities and unaffected peers. Relative differences were reported in terms of risk ratios for not achieving expected levels and were estimated using Poisson regression models with robust standard errors. Unadjusted models included only the neurodisability indicator and the outcome. As neurodisability status and educational attainment are influenced by a range of sociodemographic factors, we explored whether associations persisted after adjusting for birth month and year, neighbourhood deprivation level, region of residence, ethnicity and recorded eligibility for free school meals. We also stratified results by sex at birth to examine whether there was evidence of effect modification.

### Secondary analyses

To capture variability in educational attainment among children with neurodisability, we presented results separately for groups with frequently recorded neurodisability conditions in HES. Recorded prior to Reception, these subgroups included (1) neurodevelopmental disorders (autism, learning disability, developmental disorders); (2) cerebral palsy; (3) epilepsy; (4) Down syndrome; and (5) perinatal conditions (extremely low birth weight, extremely preterm, perinatal brain injury). The prevalence of each condition and their mortality rates are detailed in Table S3

To address the increasing number of children who remained in the Census but did not participate in primary school assessments at later time points, we conducted an additional analysis limited to those who completed all three assessments from Reception to Year 6. To assess potential selection bias, we compared the demographic and socioeconomic characteristics of children with and without neurodisability within this subgroup.

## RESULTS

Children with neurodisability had a higher mortality rate before the end of primary school (0.91% vs. 0.03% in children without neurodisabilities). The number of children alive and in the Census at the start of each school year is described in Table S4.

Of 2,351,589 pupils included in the study cohort 51,289 (2.2%) had a hospital record indicating neurodisability before starting Reception (Table S5). Children with neurodisability were more likely to be assigned male sex at birth, being Reception at age 5 instead of age 4, attend special school, be eligible for free school meals, live in more deprived neighbourhoods and be born prematurely (<37 weeks gestation) or with low birth weight (<2500g).

### Achieving expected levels in national assessments

A large proportion of children with neurodisability failed to reach expected attainment levels in every primary school assessment. Achievement rates for children with neurodisability varied from the lowest observed rate in the EYFSP assessments in Reception, where 30% met the ‘Good Level of Development’ (GLD) standard (vs. 57% of peers), to the highest observed achievement in the Reception Maths assessment, where 50% of children with neurodisability achieved expected levels (vs. 79% of peers; Table 1).

**Table 1.** Educational attainment across primary school by hospital-recorded neurodisability (ND) status.

|  | No ND before<br>school<br>n (%) | ND before<br>school<br>n (%) | Overall cohort<br>n (%) |
| --- | --- | --- | --- |
| **Total** | 2300300 (100%) | 51289 (100%) | 2351589 (100%) |
| **EYFSP** |  |  |  |
| **Good Level of Development** |  |  |  |
| Achieving expected level | 1316213 (57.2%) | 15290 (29.8%) | 1331503 (56.6%) |
| Not achieving expected level | 968942 (42.1%) | 35220 (68.7%) | 1004162 (42.7%) |
| Not assessed | 15145 (0.7%) | 779 (1.5%) | 15924 (0.7%) |
| **English** |  |  |  |
| Achieving expected level | 1584145 (68.9%) | 20229 (39.4%) | 1604374 (68.2%) |
| Not achieving expected level | 701010 (30.5%) | 30281 (59.0%) | 731291 (31.1%) |
| Not assessed | 15145 (0.7%) | 779 (1.5%) | 15924 (0.7%) |
| Standardised assessment score |  |  |  |
| Mean (SD) | 0.04 (0.96) | -0.92 (1.45) | 0.02 (0.98) |
| **Maths** |  |  |  |
| Achieving expected level | 1811241 (78.7%) | 25616 (49.9%) | 1836857 (78.1%) |
| Not achieving expected level | 473914 (20.6%) | 24894 (48.5%) | 498808 (21.2%) |
| Not assessed | 15145 (0.7%) | 779 (1.5%) | 15924 (0.7%) |
| Standardised assessment score |  |  |  |
| Mean (SD) | 0.04 (0.95) | -0.95 (1.61) | 0.02 (0.98) |
| **Key Stage 1** |  |  |  |
| **English** |  |  |  |
| Achieving expected level | 1526372 (66.4%) | 19125 (37.3%) | 1545497 (65.7%) |
| Not achieving expected level | 725270 (31.5%) | 30437 (59.3%) | 755707 (32.1%) |
| Not assessed | 48658 (2.1%) | 1727 (3.4%) | 50385 (2.1%) |
| Standardised assessment score |  |  |  |
| Mean (SD) | 0.05 (0.91) | -0.93 (1.43) | 0.03 (0.94) |
| **Maths** |  |  |  |
| Achieving expected level | 1794705 (78.0%) | 23998 (46.8%) | 1818703 (77.3%) |
| Not achieving expected level | 455723 (19.8%) | 25259 (49.2%) | 480982 (20.5%) |
| Not assessed | 49872 (2.2%) | 2032 (4.0%) | 51904 (2.2%) |
| Standardised assessment score |  |  |  |
| Mean (SD) | 0.05 (0.95) | -1.02 (1.56) | 0.02 (0.98) |
| **Key Stage 2** |  |  |  |
| **English** |  |  |  |
| Achieving expected level | 1690039 (73.5%) | 23399 (45.6%) | 1713438 (72.9%) |
| Not achieving expected level | 463019 (20.1%) | 11984 (23.4%) | 475003 (20.2%) |
| Not assessed | 147242 (6.4%) | 15906 (31.0%) | 163148 (6.9%) |
| Standardised assessment score |  |  |  |
| Mean (SD) | 0.03 (0.98) | -0.34 (1.12) | 0.02 (0.98) |
| **Maths** |  |  |  |
| Achieving expected level | 1724372 (75.0%) | 23500 (45.8%) | 1747872 (74.3%) |
| Not achieving expected level | 429608 (18.7%) | 11902 (23.2%) | 441510 (18.8%) |
| Not assessed | 146320 (6.4%) | 15887 (31.0%) | 162207 (6.9%) |
| Standardised assessment score □ |  |  |  |
| Mean (SD) □ | 0.01 (0.99) | -0.41 (1.16) | 0.00 (0.99) |

*EYFSP: Early Years Foundation Stage Profile.*

#### EYFSP: Early Years Foundation Stage Profile

While achievement rates were similar across KS1 and KS2, non-participation in assessments increased substantially. In Reception, 1.5% of children with neurodisability were not assessed under the EYFSP, compared to 0.7% of peers. At KS1, non-assessment rates rose to 3.4% in English and 4.0% in Maths (vs. 2.1% and 2.2% in peers). At KS2, non-participation in both Maths and English rose to 31%, compared to 6.4% of unaffected peers, despite continued school enrolment.

Among children with neurodisability, the substantial increase in non-participation became the primary driver of failing to reach expected levels in assessments at the end of primary school (Figure 1).

**Fig. 1.**
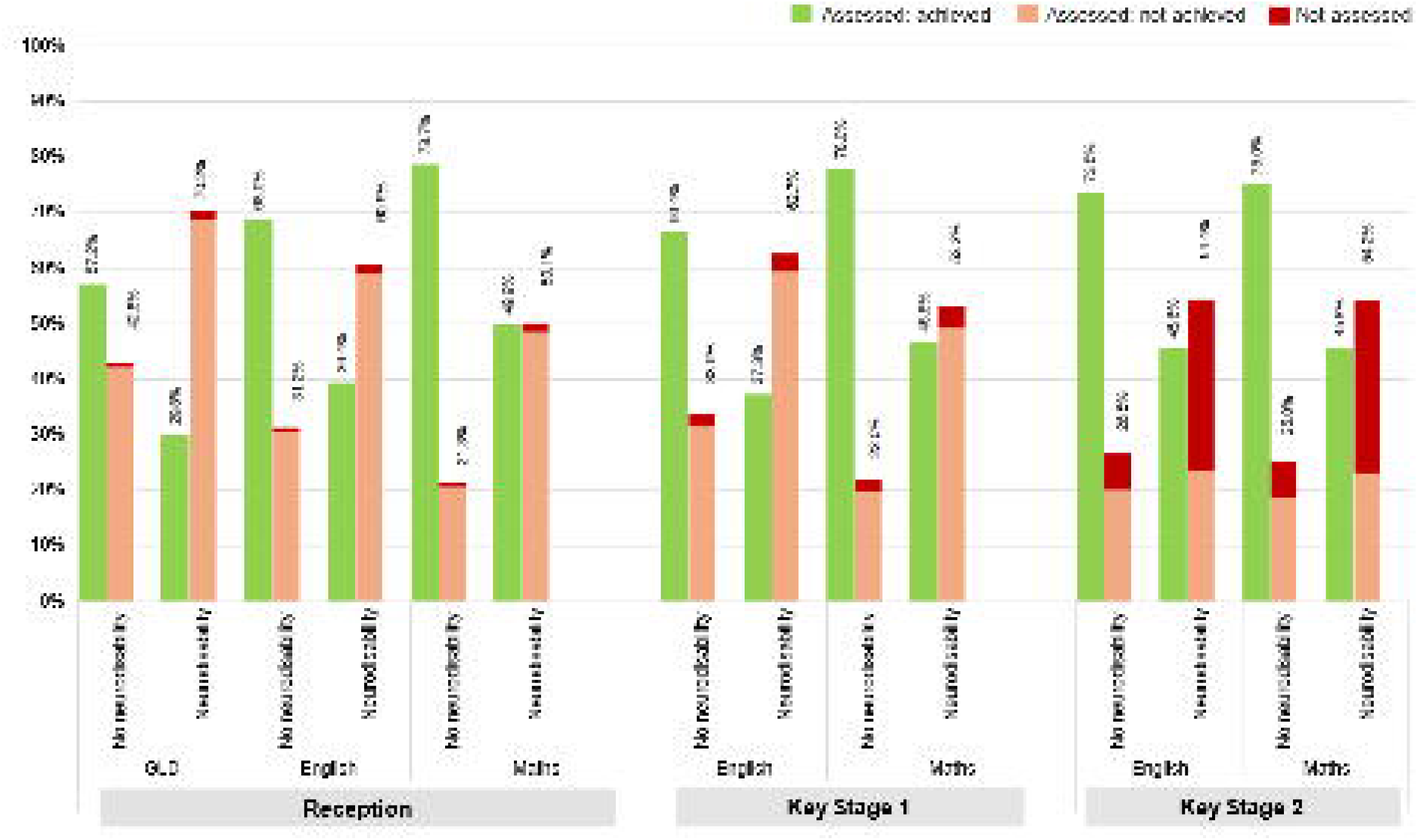

Children with neurodisability were more than twice as likely to not achieve expected levels in Maths assessments at all timepoints (Table 2). The adjusted relative risk for not achieving expected levels was 2.16 (95% CI: 2.14–2.18) at reception, 2.29 (95% CI: 2.27–2.31) at KS1, and 2.08 (95% CI: 2.07–2.10) at KS2. Similarly, children with neurodisability were over 1.7 times more likely to fail English assessments at all three stages compared to their peers. The association between neurodisability and not achieving expected levels in Maths and English was more pronounced amongst females than males (Figure 2).

**Fig. 2.**
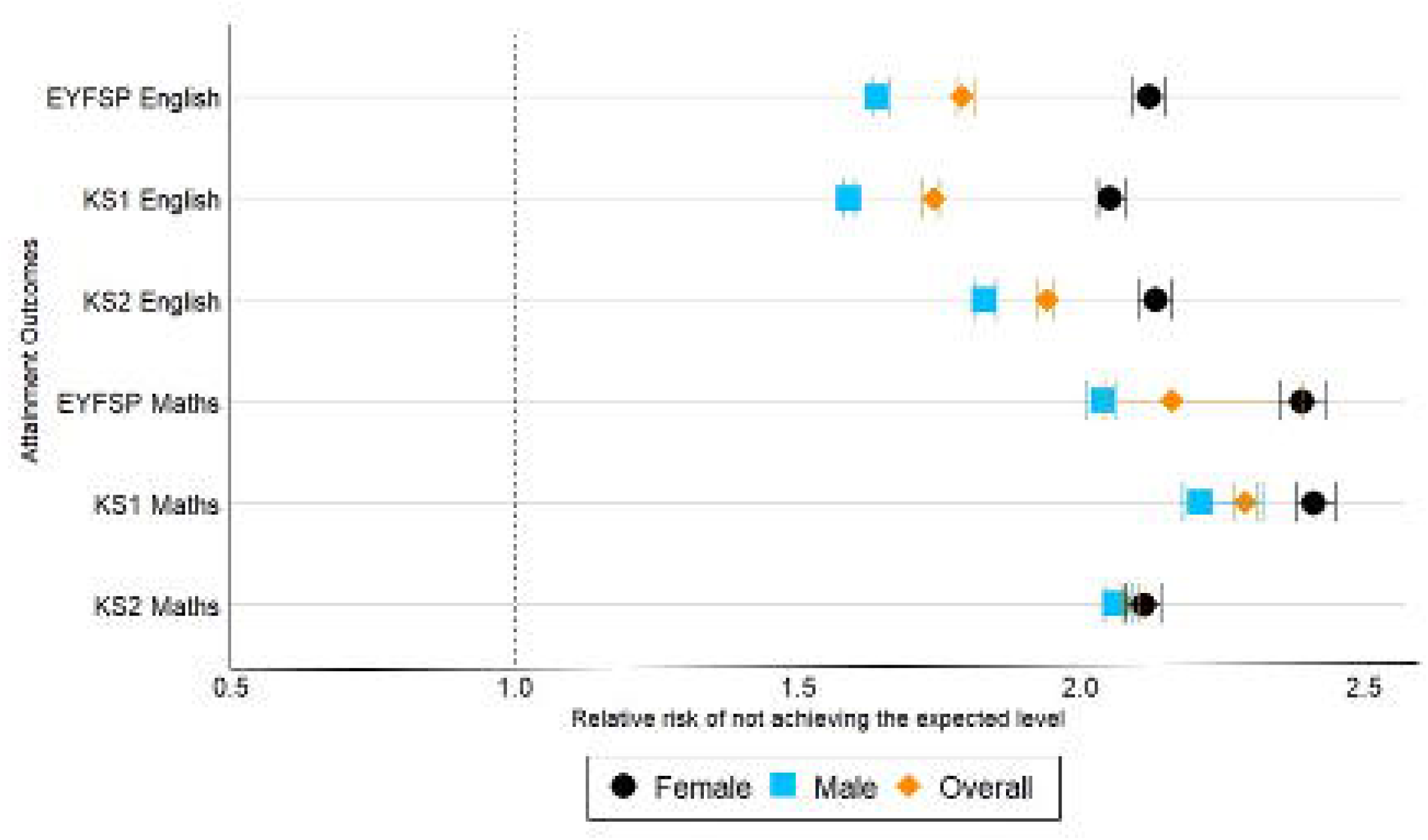

**Table 2.** Unadjusted and adjusted risk ratios for not achieving expected levels in assessments comparing children with hospital-recorded neurodisability (ND) to those without.

|  | Unadjusted |  | Adjusted |  |
| --- | --- | --- | --- | --- |
|  | No ND before school | ND before school | No ND before school | ND before school |
|  | RR (95% CI) | RR (95% CI) | RR (95% CI) | RR (95% CI) |
| **EYFSP** |  |  |  |  |
| **GLD** |  |  |  |  |
| Overall | 1 (Ref) | 1.64 (1.63 – 1.65) | 1 (Ref) | 1.54 (1.53 – 1.55) |
| Males | 1 (Ref) | 1.46 (1.45 – 1.47) | 1 (Ref) | 1.42 (1.41 – 1.43) |
| Females | 1 (Ref) | 1.87 (1.85 – 1.89) | 1 (Ref) | 1.79 (1.78 – 1.81) |
| **English** |  |  |  |  |
| Overall | 1 (Ref) | 1.94 (1.93 – 1.96) | 1 (Ref) | 1.79 (1.78 – 1.81) |
| Males | 1 (Ref) | 1.72 (1.70 – 1.73) | 1 (Ref) | 1.64 (1.63 – 1.66) |
| Females | 1 (Ref) | 2.26 (2.26 – 2.28) | 1 (Ref) | 2.12 (2.09 – 2.15) |
| **Maths** |  |  |  |  |
| Overall | 1 (Ref) | 2.35 (2.33 – 2.38) | 1 (Ref) | 2.16 (2.14 – 2.18) |
| Males | 1 (Ref) | 2.17 (2.14 – 2.19) | 1 (Ref) | 2.04 (2.01 – 2.06) |
| Females | 1 (Ref) | 2.58 (2.53 – 2.61) | 1 (Ref) | 2.39 (2.35 – 2.43) |
| **Key Stage 1** |  |  |  |  |
| **English** |  |  |  |  |
| Overall | 1 (Ref) | 1.86 (1.85 – 1.88) | 1 (Ref) | 1.74 (1.72 – 1.75) |
| Males | 1 (Ref) | 1.65 (1.63 – 1.66) | 1 (Ref) | 1.59 (1.58 – 1.60) |
| Females | 1 (Ref) | 2.16 (2.14 – 2.19) | 1 (Ref) | 2.05 (2.03 – 2.08) |
| **Maths** |  |  |  |  |
| Overall | 1 (Ref) | 2.42 (2.40 – 2.44) | 1 (Ref) | 2.29 (2.27 – 2.31) |
| Males | 1 (Ref) | 2.31 (2.28 – 2.33) | 1 (Ref) | 2.21 (2.18 – 2.32) |
| Females | 1 (Ref) | 2.54 (2.51 – 2.58) | 1 (Ref) | 2.41 (2.38 – 2.45) |
| **Key Stage 2** |  |  |  |  |
| **English** |  |  |  |  |
| Overall | 1 (Ref) | 2.05 (2.03 – 2.07) | 1 (Ref) | 1.94 (1.92 – 1.95) |
| Males | 1 (Ref) | 1.90 (1.88 – 1.92) | 1 (Ref) | 1.83 (1.81 – 1.85) |
| Females | 1 (Ref) | 2.22 (2.19 – 2.25) | 1 (Ref) | 2.13 (2.10 – 2.16) |
| **Maths** |  |  |  |  |
| Overall | 1 (Ref) | 2.16 (2.15 – 2.18) | 1 (Ref) | 2.08 (2.07 – 2.10) |
| □ □ □ □ |  |  |  |  |
| Males □ □ | 1 (Ref) □ | 2.14 (2.12 – 2.16) | 1 (Ref) □ | □ 2.06 (2.04 – 2.09) |
| □ □ □ □ |  |  |  |  |
| Females □ □ | 1 (Ref) □ | 2.19 (2.16 – 2.22) | 1 (Ref) □ | □ 2.11 (2.08 – 2.14) |

*RR = Risk Ratio. CI = Confidence Interval. Adjusted models controlled for sex at birth, birth month, year of birth, neighbourhood deprivation level, region of residence, ethnicity and recorded eligibility for free school meals.*

### Secondary analysis

Educational attainment varied considerably among children with different neurodisability-associated conditions (Table S6). In Maths assessments throughout primary school, attainment was highest among children with perinatal conditions, with 55–60% achieving expected levels. This was followed by children with epilepsy (34–38%), cerebral palsy (27–36%), and neurodevelopmental disorders (24–25%), while the lowest attainment was observed in children with Down syndrome (1–4%; Figure 3). A similar pattern was observed in English assessments (Figure S3).

**Fig. 3.**
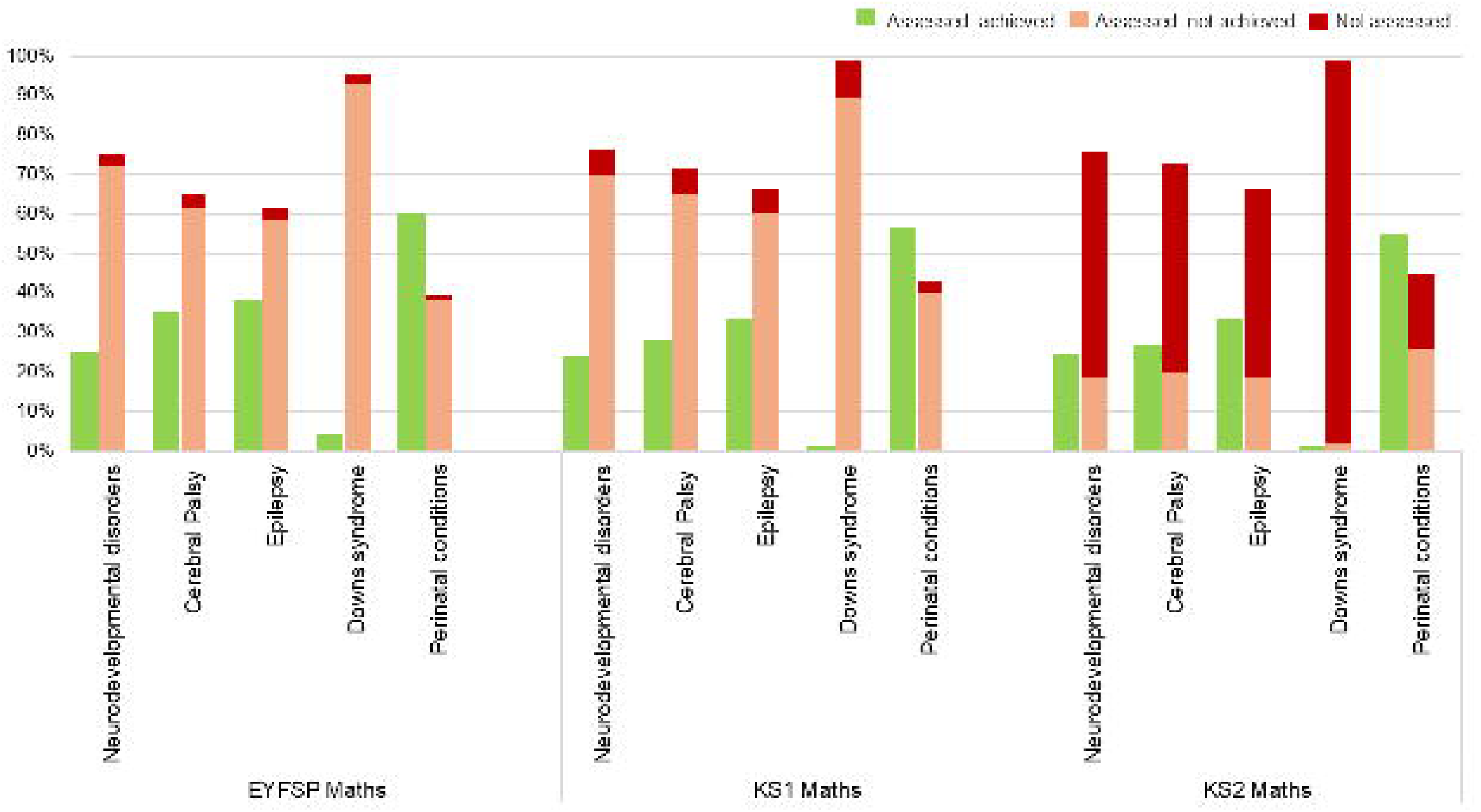

Patterns of non-participation followed those seen in the overall neurodisability group, increasing by KS2. Notably, 97% of children with Down syndrome were not assessed in KS2, representing the highest rate of non-participation, whereas 19% of children with perinatal conditions were not assessed, indicating the highest participation rate among the neurodisability subgroups (Figure 3).

Of the 51,289 children with neurodisability in our study, 34,848 (67.9%) completed all three assessments across primary school in both Maths and English (vs. 91.9% of peers). Children with neurodisability who completed all three assessments were more likely to meet expected levels at EYFSP, KS1 and KS2 compared with children with neurodisability who missed at least one assessment (Table S7). However, this group were still less likely to achieve expected levels than their unaffected peers.

## DISCUSSION

Our study of more than 2 million children in England showed that children with neurodisability consistently did not achieve expected levels of attainment in primary school assessments, with fewer than half of these children meeting national standards across all Maths and English assessments. Children with neurodisability were over twice as likely to fail all Maths assessments and 1.7 times more likely to fail all English assessments. Non-participation in assessments increased substantially between KS1 and KS2, becoming the main reason for not achieving expected levels of attainment at the end of primary school. The risk of not achieving expected levels in primary school assessments was more pronounced when comparing females with neurodisabilities to their unaffected peers. This disparity may stem from differences in how neurodisabilities manifest in females, where symptoms are often less overt, leading to underdiagnosis in milder cases.^19^ As a result, the diagnosed female neurodisability group may represent a more severely affected population with a higher need for additional support.

Attainment outcomes varied significantly by neurodisability subgroup. Children with perinatal conditions had the best attainment outcomes among children with neurodisability, with over 55% of them achieving expected levels in all primary school Maths assessments. The lowest attainment was observed among children with Down syndrome, where under 5% of children achieved expected levels every primary school assessment.

Our findings provide novel prognostic insights into the educational journey of children with neurodisability. This has not previously been investigated on a population scale in England. A key strength of our study was the use of linked health and education data encompassing an entire cohort of children born in England between 2003 and 2008. The large size and extensive population coverage of the Hospital Episode Statistics (HES) and National Pupil Database (NPD) allowed us to examine outcomes for pupils with relatively rare exposures, including subgroups of neurodisability, and make comparisons with a high degree of precision. Additionally, more than 95% of children enrolled in reception remained enrolled in Year 6, enabling the longitudinal follow up of children with and without a recorded neurodisability throughout primary school for nearly the whole population.

Our study has several limitations. Case ascertainment relied on applying phenotype code lists deterministically to diagnosis and procedure codes, which are primarily recorded for the financial reimbursement of medical care. As a result, conditions that are not managed in secondary care may be underrepresented, since children with these conditions are less likely to have a hospital admission (for example, if children receive much of their care from community paediatrics, mental health services or primary care). The underrepresented conditions include autism spectrum disorder, conduct disorder, hyperkinetic disorders and perinatal brain injury.^3^ Thus, our population are more likely to represent a group with more complex health needs and children with milder forms of neurodisability may be misclassified into the unaffected group, potentially underestimating the true differences in attainment between children with and without a recorded neurodisability.

Another limitation of our study is the variation in scoring methods across the three primary school assessments. Both the EYFSP and KS1 assessments are teacher-assessed, whereas KS2 assessments are externally marked. This variation may limit direct comparability and introduce bias in assessment outcomes, as differences in teachers’ perceptions of a child’s abilities, as well as school-level marking practices, could influence how assessments are scored. However, these remain the only primary school outcome measures available in the NPD. Additionally, to account for national changes in assessment structures during period of our study (e.g., changes to EYFSP structure in 2012),^20^ it was important that we applied standardisation rules to ensure consistent measurement across years. As no established standard exists for this process, we made methodological decisions to maximise comparability (Table S1).

### Our findings indicate that national standardised assessments are not a meaningful measure of educational progress for many children with neurodisability

These assessments capture only a narrow aspect of a child’s development and success, overlooking more functional and granular measures of progress. Over 30% of children with neurodisability do not participate in KS2 assessments, meaning they are officially recorded as not achieving expected attainment levels despite potentially making significant progress in areas not measured by national assessments. In state-funded schools in England, children working below national curriculum standards are typically assessed under the Engagement Model,^21^ which takes a child-centered approach, focusing on abilities rather than disabilities. This model tracks progress across five key areas: exploration, anticipation, realisation, persistence, and initiation, recognising the complex interplay of physical, sensory, communication, and learning disabilities in shaping a child’s educational progress. Unlike standardised assessments, the Engagement Model provides a more personalised and meaningful measure of progress. However, schools in England are not required to report Engagement Model data to the Department for Education, meaning it is absent from the National Pupil Database (NPD) and cannot be evaluated. Integrating these outcomes into national datasets is essential for tracking long-term educational progress in children with neurodisability on a population-level.

### We demonstrate that children who are likely to face functional impairments from neurodisability can be identified in hospital records before school entry, presenting a valuable opportunity for early intervention

Given these children are already known to health services, there is clear potential for better coordination between health and education to ensure that pre-identified support needs are in place upon starting school. Only a third of children with neurodisability were deemed ‘school ready’ by the Good Level of Development (GLD) at the end of Reception, yet nearly all (99%, Table S4) transitioned to Year 1 the following term. Children with neurodisability would therefore benefit from additional educational support during this transition, where health and education services could collaborate to prioritise the assessment of children with neurodisability for Special Educational Needs and Disability (SEND) provision. While a large proportion (75%) of children with neurodisability in England finish primary school having received SEN provision at some point, this is not always provided in a timely manner, if at all.^3^ A more integrated national health and education system could enable earlier assessments, ensuring children receive the educational support they require as soon as it is needed.

### Education is a key social determinant of outcomes throughout adulthood

Thus, further research is needed to follow these children through secondary school and beyond, as children with neurodisability are more likely to leave school before 16, face exclusion and be not in education, employment, or training (NEET).^5, 22, 23, 24^

Our study adds to a limited body of evidence on neurodisability in England. We demonstrate that children with neurodisability are less likely to achieve expected levels of attainment in primary school compared to their peers. The progress made by these children are currently inadequately captured in population-level data.

Supporting children with chronic conditions such as neurodisability will help to improve their participation in society and transition to independence.^25^

## Data Availability

The ECHILD database is made available for free for approved research based in the UK, via the ONS Secure Research Service. Enquiries to access the ECHILD database can be made by emailing. Researchers will need to be approved and submit a successful application to the ECHILD Data Access Committee and ONS Research Accreditation Panel to access the data with strict statistical disclosure controls of all outputs of analyses.

## Acknowledgments

We would like to acknowledge all children, young people, and families whose de-identified data are used in this research. We would also like to acknowledge the contribution of the wider HOPE Study Group to this work: Sarah Barnes, Kate Boddy, Kristine Black-Hawkins, Lorraine Dearden, Bianca De Stavola, Johnny Downs, Martin Doyle, William Farr, Tamsin Ford, Lucy Karwatowska, Kate Lewis, Matthew Lilliman, Stuart Logan, Vincent Nguyen, Jugnoo Rahi, Jennifer Saxton, Joachim Tan, and Isaac Winterburn. We thank Ruth Blackburn, Matthew Jay, Farzan Ramzan, and Tony Stone for ECHILD database support.

## Ethics Approval

Permissions to use linked, de-identified data from Hospital Episode Statistics and the National Pupil Database were granted by the Department for Education (DR200604.02B) and NHS Digital (DARS-NIC-381972). Ethical approval for the ECHILD project was granted by the National Research Ethics Service (17/LO/1494), NHS Health Research Authority Research Ethics Committee (20/EE/0180) and UCL Great Ormond Street Institute of Child Health’s Joint Research and Development Office (20PE06).

## Data Availability Statement

The ECHILD database is made available for free for approved research based in the UK, via the ONS Secure Research Service. Enquiries to access the ECHILD database can be made by. Researchers will need to be approved and submit a successful application to the ECHILD Data Access Committee and ONS Research Accreditation Panel to access the data, with strict statistical disclosure controls of all outputs of analyses.

