## Supplementary Materials for "Educational attainment among primary school children with neurodisability: A population-based cohort study using linked education and health data from England"

A) **Deviations from the study protocol**

A protocol for this study was published in March 2024.^1^ Changes to the analysis plan are detailed below:

1. We planned to use children's hospital admission and mortality records from birth until August 31st of Year 6 to identify neurodisability and to stratify our analyses based on the timing of the first hospital-recorded neurodisability (before school vs. during school). However, further validation of the code list and the recording of neurodisability in Hospital Episode Statistics (HES) revealed that these two groups represent children with different types of conditions rather than delayed diagnoses^2^. As a result, we chose to focus on children whose neurodisability was first recorded before school entry, as they are already known to healthcare services prior to starting school.
2. We chose to focus on five subgroups of neurodisability, selecting the most common ones within our cohort. These were: 1) neurodevelopmental disorders; 2) epilepsy, 3) cerebral palsy; 4) Down syndrome; 5) perinatal conditions.
3. We ultimately decided not to use linear mixed modeling to predict attainment trajectories as, by the later Key Stages (Year 2 and Year 6), a significant number of children did not sit the exams, and this missing data could not be assumed to be random. As a result, the model failed to provide meaningful or interpretable results. Our primary goal was to generate prognostic and descriptive insights at the population level, rather than to numerically estimate trajectories using a method that was not well-suited to the nature of our data. We aimed to produce valuable, practical information that could be easily understood and interpreted by teachers, caregivers, and healthcare professionals, rather than relying on a statistical approach that did not align with the reality of the data.
4. In the protocol we proposed to investigate the effect of sociodemographic variables and other potential confounders between neurodisability and attainment outcomes. This was outside the scope of this paper and has been reserved for more in-depth and focused future work on inequalities. In our regression models, we adjusted for sociodemographic variables to quantify whether differences in attainment were attenuated once these characteristics were accounted for.
B) **Deviations of study cohort and measures**

**Cohort derivation**

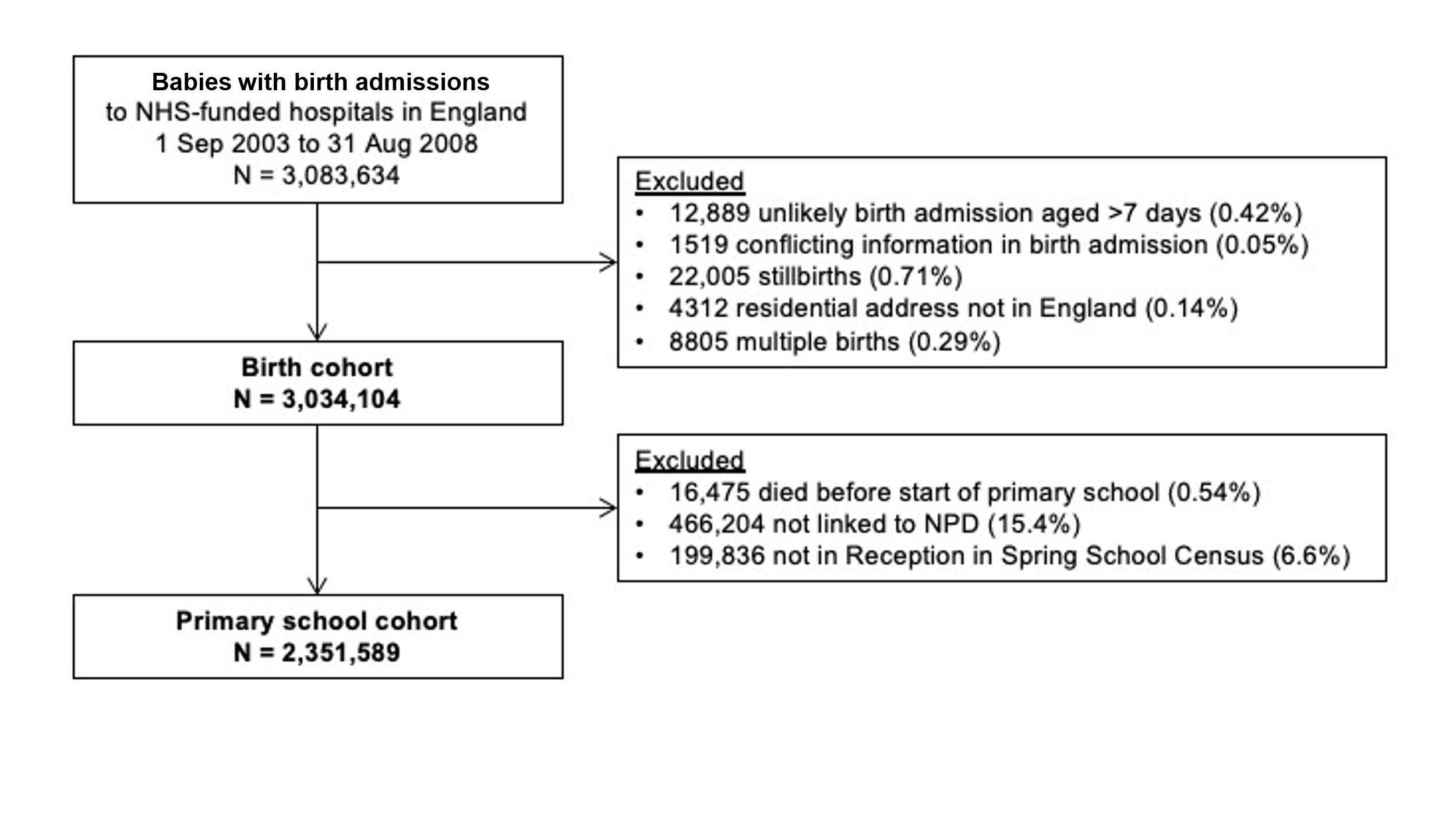
**Figure S1.** Construction of primary school cohort.

We identified all potential birth admissions in HES between 1 Sep 2003 and 31 Aug 2008 (3,083,643). Children born from 1st September 2003 are the first complete academic year cohorts born after the introduction of the NHS Number for Babies (NN4B) which improved linkage rates between birth admissions and subsequent admissions.^3^ These cohorts are all also expected to finish primary school before the start of the COVID-19 pandemic in March 2020. We focus on singleton births due to the challenge of accurately linking records for twins. The birth cohort of singleton children consisted of 3,034,104 children (98.4% of the birth admissions initially identified). Of these, 2,351,589 (83.4%) were enrolled in Reception at state-funded primary school (aged 4/5).

We included children enrolled in Reception (age 4/5) at state-funded mainstream and special schools in the January (Spring) School Census. While mandated primary school begins in Year 1 (age 5/6) in England, most children are enrolled in Reception. We focus on the Spring School Census, as it is used to allocate school funding, and so is believed to be the most complete of the three School Censuses conducted every year (Autumn, Spring and Summer). Children who were recorded as enrolled in Reception two or more years outside of their expected school start year given their birth date were excluded. Please see the HOPE study umbrella protocol for more information.^4^

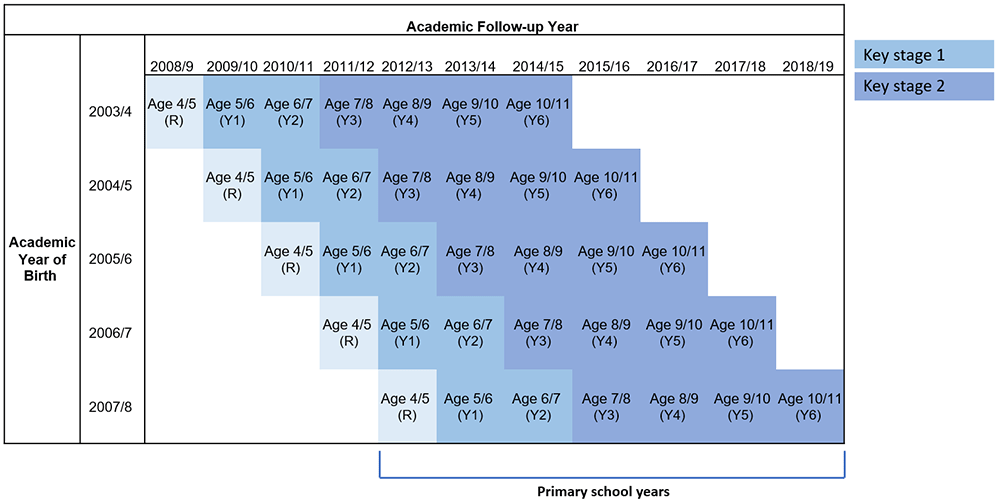
**Figure S2.** Timeline for follow up of primary school cohort.

*R = reception; Y = year; birth and follow-up year defined according to the academic calendar (e.g. 2003/4 includes 1st September 2003 to 31st August 2004, inclusive).*

**Deriving outcome measures**

**Table S1.** Educational attainment outcome measures from the NPD.

| **Subject** | **Years** | **Variables in NPD^5^** |
| --- | --- | --- |
| **EYFSP** | | |
| GLD | 2002/3-2011/12 | FSP_EYFSP_TOTAL |
| Maths |  | FSP_PSRN_TOTAL |
| English |  | FSP_CLL_TOTAL |
| GLD | 2012/13 onwards | Sum of:  COM_G01, COM_G02, COM_G03, PHYG04, PHY_G05 PSE_G06, PSE_G07, PSE_G08,  LIT_G09, LIT_G10, MAT_G11, MAT_G12,  UTW_G13, UTW_G14, UTW_G15 EXP_G16, EXP_G17 |
| Maths |  | MAT_G11, MAT_G12 |
| English |  | COM_G01, COM_G02, COM_G03, LIT_G09, LIT_G10 |
| **Key Stage 1** | | |
| Maths | 2002/3-2014/15 | KS1_MATHS,  KS1_MAT_LEV |
| English |  | KS1_READING, KS1_WRIT, KS1_READ_LEV,  KS1_WRIT_LEV |
| Maths | 2015/16 onwards | KS1_MATH_OUTCOME |
| English |  | KS1_EXPECT_LEV_READ  KS1_EXPECT_LEV_WRIT |
| **Key Stage 2** | | |
| Maths | 2002/3-2014/15 | KS2_MATTOMRK, KS2_MATLEV |
| English |  | KS2_READLEV, KS2_READTOMRK |
| Maths | 2015/16 onwards | KS2_MATMRK, KS2_MATEXP |
| English |  | KS2_READMRK, KS2_READEXP |

*Multiple Imputation conducted on missing data in 2009/10 due to industrial action. ECHILD “How To Guide – Guide 9” contains more information:* [*https://howto.echild.ac.uk/guide09_exam*](https://howto.echild.ac.uk/guide09_exam)

**Pupil characteristics**

**Table S2.** Pupil characteristics extracted from ECHILD.

| **Variable** | **Description** | **Dataset** | **Included in regressions** |
| --- | --- | --- | --- |
| Sex at birth | Based on sex recorded in HES birth admission (male/female) | HES | Yes |
| Age at school start | Based on birth date in HES and year when child is recorded as enrolled in Reception in the Spring Census (or as ‘not following the national curriculum’ in the year they turned 5). | HES/  NPD | No |
| Birth month | Based on date of birth recorded in HES birth admission; regrouped as September-October-November, December-January-February, March-April-May, and June-July-August. | HES | Yes |
| Academic year of birth | Based on date of birth in HES birth admission. Pupils are grouped according to which academic year (from 1^st^ September to 31^st^ August the following year) they were born in. This variable therefore represents the school cohort a pupil is part of. | HES | Yes |
| IDACI quintile | Taken from earliest recording in the School Census (usually Reception, or Year 1). IDACI is an area-level measure of the proportion of children under the age of 16 living in low-income households.^6^ We used IDACI quintile associated with a pupil’s home address and retain a category for missing IDACI. | NPD | Yes |
| Recorded free school meals | Taken from earliest recording in School Census (usually Reception, or Year 1). We use free school meals (FSM) as a proxy for household socioeconomic position, since a child’s eligibility to receive FSM depends on whether their parents or guardians are in receipt of benefits. | NPD | Yes |
| Region of residence | Taken from earliest recording in School Census (usually Reception or Year 1). Using the local authority of a pupil’s recorded home address, we create groups corresponding to Government Office Region (GOR). North East, North West, Yorkshire and the Humber, East Midlands, West Midlands, East of England, London, South East, South West. We retain a category for missing GOR. Categorical variable. | NPD | Yes |
| Ethnicity | Mode of ethnicity recorded across any School Census that the pupil was included in. We regroup ethnicity into the following categories: Asian or Chinese, Black, Mixed, White, Any Other Ethnic Group, and Unclassified/Missing. Categorical variable. | NPD | Yes |
| Birth weight  Gestational age  Maternal age | Birth characteristics were obtained from birth admission (following data cleaning described elsewhere)^3^ and completed using information from the linked maternal record where available.^7^ | HES | No |
| Special school attendance | Binary indicator of ever attending special school based on school information recorded in NPD school census and Get Information About Schools Dataset (GIAS). We derived an indicator of whether a child had ever been enrolled in a special school by end of Year 6 (age 10/11). | NPD/  GIAS | No |
| Death during the study period | Binary indicator based on linked ONS mortality records and information about in-hospital deaths from HES (data cleaning based on methods described previously).^3^ | ONS | No |

*ECHILD: Education and Child Health Insights from Linked Data. HES: Health Episode Statistics Admitted Patient care. IDACI: Index of Deprivation Affecting Children Index. ONS: Office for National Statistics. NPD: National Pupil Database.*

**Table S3.** Prevalence and mortality rates of neurodisability subgroups.

|  | **Male** | **Female** | **Overall** | **Mortality rate** |
| --- | --- | --- | --- | --- |
|  | **n (%)** | **n (%)** | **n (%)** | **n (%)** |
| Total | 1207687 (100%) | 1143902 (100%) | 2351589 (100%) | 915 (0.04%) |
| Any neurodisability | 29856 (2.5%) | 21433 (1.9%) | 51289 (2.2%) | 466 (0.91%) |
| NDD | 6073 (0.5%) | 2800 (0.2%) | 8873 (0.4%) | 203 (2.29%) |
| Cerebral palsy | 2627 (0.2%) | 1887 (0.2%) | 4514 (0.2%) | 174 (3.85%) |
| Epilepsy | 4074 (0.3%) | 3130 (0.3%) | 7204 (0.3%) | 233 (3.23%) |
| Down syndrome | 1333 (0.1%) | 1049 (0.1%) | 2382 (0.1%) | 11 (0.46%) |
| Perinatal conditions | 9792 (0.8%) | 7965 (0.7%) | 17757 (0.8%) | 102 (0.57%) |

*Neurodisability subtypes recorded in hospital data before start of reception (age 4/5). NDD = Neurodevelopmental conditions. Mortality rate by Year 6.*

**Loss to follow-up**

**Table S4.** Number and percentage of cohort members alive on 1^st^ September of each school year and enrolled by school year and neurodisability status.

| **School Year** | **Alive 1^st^ Sep (% of total cohort)** | **Enrolled**  **(% of alive)** | **% of enrolled with missing absence data** |
| --- | --- | --- | --- |
| ***ND*** |  |  |  |
| R | 51289 (100%) | 51289 (100%) | - |
| Y1 | 51227 (99.9%) | 50939 (99%) | 0.5% |
| Y2 | 51137 (99.7%) | 50555 (99%) | 0.4% |
| Y3 | 51081 (99.6%) | 50144 (98%) | 0.4% |
| Y4 | 51027 (99.5%) | 49833 (98%) | 0.4% |
| Y5 | 50963 (99.4%) | 49463 (97%) | 0.4% |
| Y6 | 50908 (99.3%) | 49118 (96%) | 0.8% |
| ***No ND*** |  |  |  |
| R | 2300300 (100%) | 2300300 (100%) | - |
| Y1 | 2300241 (>99.9%) | 2285362 (99%) | 0.1% |
| Y2 | 2300153 (>99.9%) | 2268905 (99%) | 0.1% |
| Y3 | 2300054 (>99.9%) | 2248624 (98%) | 0.1% |
| Y4 | 2299962 (>99.9%) | 2234572 (97%) | 0.1% |
| Y5 | 2299865 (>99.9%) | 2220989 (97%) | 0.1% |
| Y6 | 2299766 (>99.9%) | 2208427 (96%) | 0.1% |

*ND = neurodisability recorded in hospital data before start of Reception. R = Reception year.*

**Table S5.** Characteristics of the study cohort by hospital-recorded neurodisability (ND) status. Includes singleton children born in NHS-funded hospitals in England between 01/09/2003 and 31/08/2008, linked to NPD and enrolled in Reception (age 4/5) at state-funded schools in England.

|  | **No ND before school**  **n (%)** | **ND before school**  **n (%)** | **ND prevalence in group**  **%** |
| --- | --- | --- | --- |
| **Total** | 2300300 (100.0%) | 51289 (100.0%) | 2.23% |
| **Sex at birth** |  |  |  |
| Male | 1177831 (51.2%) | 29856 (58.2%) | 2.53% |
| Female | 1122469 (48.8%) | 21433 (41.8%) | 1.91% |
| **Age at school start** |  |  |  |
| Age 4 | 2299368 (100.0%) | 50969 (99.4%) | 2.22% |
| Age 5 | 932 (0.0%) | 320 (0.6%) | 34.3% |
| **Ever in special school (R-Y6)** |  |  |  |
| No | 2283059 (99.3%) | 42117 (82.1%) | 1.84% |
| Yes | 17241 (0.7%) | 9172 (17.9%) | 53.2% |
| **Died before end of Year 6** |  |  |  |
| No | 2299605 (99.9%) | 50823 (99.1%) | 2.23% |
| Yes | 695 (0.03%) | 466 (0.9%) | 67.1% |
| **Month of birth** |  |  |  |
| Sep-Oct-Nov | 576753 (25.1%) | 13224 (25.8%) | 2.29% |
| Dec-Jan-Feb | 548324 (23.8%) | 12384 (24.1%) | 2.26% |
| Mar-Apr-May | 573943 (25.0%) | 12854 (25.1%) | 2.24% |
| Jun-Jul-Aug | 601280 (26.1%) | 12827 (25.0%) | 2.13% |
| **Academic year of birth** |  |  |  |
| 2003/4 | 426352 (18.5%) | 9326 (18.2%) | 2.19% |
| 2004/5 | 451075 (19.6%) | 9716 (18.9%) | 2.15% |
| 2005/6 | 465539 (20.2%) | 10227 (19.9%) | 2.20% |
| 2006/7 | 476322 (20.7%) | 10644 (20.8%) | 2.23% |
| 2007/8 | 481012 (20.9%) | 11376 (22.2%) | 2.37% |
| **Ethnic group** |  |  |  |
| Any Other Ethnic Group | 27252 (1.2%) | 544 (1.1%) | 2.00% |
| Asian or Chinese | 203546 (8.8%) | 4786 (9.3%) | 2.35% |
| Black | 105932 (4.6%) | 2638 (5.1%) | 2.49% |
| Mixed | 107799 (4.7%) | 2425 (4.7%) | 2.25% |
| White | 1630402 (70.9%) | 35585 (69.4%) | 2.18% |
| Missing | 225369 (9.8%) | 5311 (10.4%) | 2.36% |
| **Free school meals** **eligible** |  |  |  |
| No | 1883776 (81.9%) | 38614 (75.3%) | 2.05% |
| Yes | 416524 (18.1%) | 12675 (24.7%) | 3.04% |
| **IDACI quintile** |  |  |  |
| 1 (most deprived) | 602297 (26.2%) | 15731 (30.7%) | 2.61% |
| 2 | 479402 (20.8%) | 11298 (22.0%) | 2.36% |
| 3 | 423113 (18.4%) | 9098 (17.7%) | 2.15% |
| 4 | 403952 (17.6%) | 7908 (15.4%) | 1.96% |
| 5 (least deprived) | 383741 (16.7%) | 7045 (13.7%) | 1.84% |
| Missing | 7795 (0.3%) | 209 (0.4%) | 2.68% |
| **Region of residence** |  |  |  |
| East Midlands | 200314 (8.7%) | 4586 (8.9%) | 2.29% |
| East of England | 253010 (11.0%) | 4862 (9.5%) | 1.92% |
| London | 351658 (15.3%) | 7559 (14.7%) | 2.15% |
| North East | 118240 (5.1%) | 2920 (5.7%) | 2.47% |
| North West | 300798 (13.1%) | 7243 (14.1%) | 2.41% |
| South East | 362062 (15.7%) | 7843 (15.3%) | 2.17% |
| South West | 220839 (9.6%) | 5167 (10.1%) | 2.34% |
| West Midlands | 236285 (10.3%) | 5598 (10.9%) | 2.37% |
| Yorkshire and the Humber | 249510 (10.8%) | 5311 (10.4%) | 2.15% |
| Missing | 7584 (0.3%) | 200 (0.4%) | 2.64% |
| **Gestational age** |  |  |  |
| <32 weeks | 7738 (0.3%) | 4914 (9.6%) | 63.5% |
| 32-36 weeks | 76674 (3.3%) | 3430 (6.7%) | 4.47% |
| 37-41 weeks | 1373684 (59.7%) | 23468 (45.8%) | 1.71% |
| 42+ weeks | 70196 (3.1%) | 1139 (2.2%) | 1.62% |
| Missing | 772008 (33.6%) | 18338 (35.8%) | 2.38% |
| **Birthweight** |  |  |  |
| <2500 grams | 94503 (4.1%) | 10420 (20.3%) | 11.0% |
| 2500-3999.99 grams | 1473225 (64.0%) | 25058 (48.9%) | 1.70% |
| 4000+ grams | 204521 (8.9%) | 3063 (6.0%) | 1.50% |
| Missing | 528051 (23.0%) | 12721 (24.8%) | 2.41% |
| **Maternal age** |  |  |  |
| <20 years | 157179 (6.8%) | 4047 (7.9%) | 2.57% |
| 20-29 years | 1021078 (44.4%) | 22357 (43.6%) | 2.19% |
| 30-34 years | 618152 (26.9%) | 12069 (23.5%) | 1.95% |
| 35-39 years | 344482 (15.0%) | 7547 (14.7%) | 2.19% |
| 40+ years | 70760 (3.1%) | 2040 (4.0%) | 2.88% |
| Missing | 88649 (3.9%) | 3229 (6.3%) | 3.64% |

*IDACI: Income Deprivation Affecting Children Index*

C) **Additional results**

**Table S6.** Educational attainment outcomes across primary school assessments by neurodisability subgroup.

|  | **NDD**  **n (%)** | **Cerebral Palsy**  **n (%)** | **Epilepsy**  **n (%)** | **Down Syndrome**  **n (%)** | **Perinatal**  **n (%)** |
| --- | --- | --- | --- | --- | --- |
| **Total** | 8873 (100%) | 4514 (100%) | 7204 (100%) | 2382 (100%) | 17757 (100%) |
| **EYFSP** |  |  |  |  |  |
| ***GLD*** |  |  |  |  |  |
| Achieving expected level | 922 (10.4%) | 682 (15.1%) | 1558 (21.6%) | 23 (1.0%) | 6725 (37.9%) |
| Not achieving expected level | 7701 (86.8%) | 3679 (81.5%) | 5426 (75.3%) | 2299 (96.5%) | 10822 (60.9%) |
| Not assessed | 250 (2.8%) | 153 (3.4%) | 220 (3.1%) | 60 (2.5%) | 210 (1.2%) |
| ***English*** |  |  |  |  |  |
| Achieving expected level | 1481 (16.7%) | 1143 (25.3%) | 2131 (29.6%) | 70 (2.9%) | 8708 (49.0%) |
| Not achieving expected level | 7142 (80.5%) | 3218 (71.3%) | 4853 (67.4%) | 2252 (94.5%) | 8839 (49.8%) |
| Not assessed | 250 (2.8%) | 153 (3.4%) | 220 (3.1%) | 60 (2.5%) | 210 (1.2%) |
| Standardised assessment score |  |  |  |  |  |
| Mean (SD) | -1.82 (1.43) | -1.56 (1.56) | -1.43 (1.61) | -2.73 (0.97) | -0.54 (1.28) |
| ***Maths*** |  |  |  |  |  |
| Achieving expected level | 2228 (25.1%) | 1588 (35.2%) | 2756 (38.3%) | 102 (4.3%) | 10694 (60.2%) |
| Not achieving expected level | 6395 (72.1%) | 2773 (61.4%) | 4228 (58.7%) | 2220 (93.2%) | 6853 (38.6%) |
| Not assessed | 250 (2.8%) | 153 (3.4%) | 220 (3.1%) | 60 (2.5%) | 210 (1.2%) |
| Standardised assessment score |  |  |  |  |  |
| Mean (SD) | -1.87 (1.72) | -1.65 (1.85) | -1.53 (1.85) | -3.10 (1.30) | -0.55 (1.38) |
| **Key Stage 1** |  |  |  |  |  |
| ***English*** |  |  |  |  |  |
| Achieving expected level | 1455 (16.4%) | 960 (21.3%) | 1872 (26.0%) | 27 (1.1%) | 8267 (46.6%) |
| Not achieving expected level | 6978 (78.6%) | 3293 (73.0%) | 4975 (69.1%) | 2221 (93.2%) | 8949 (50.4%) |
| Not assessed | 440 (5.0%) | 261 (5.8%) | 357 (5.0%) | 134 (5.6%) | 541 (3.0%) |
| Standardised assessment score |  |  |  |  |  |
| Mean (SD) | -1.84 (1.39) | -1.60 (1.46) | -1.47 (1.51) | -2.90 (0.63) | -0.53 (1.25) |
| ***Maths*** |  |  |  |  |  |
| Achieving expected level | 2122 (23.9%) | 1268 (28.1%) | 2422 (33.6%) | 31 (1.3%) | 10078 (56.8%) |
| Not achieving expected level | 6221 (70.1%) | 2931 (64.9%) | 4357 (60.5%) | 2136 (89.7%) | 7094 (40.0%) |
| Not assessed | 530 (6.0%) | 315 (7.0%) | 425 (5.9%) | 215 (9.0%) | 585 (3.3%) |
| Standardised assessment score |  |  |  |  |  |
| Mean (SD) | -1.98 (1.59) | -1.77 (1.61) | -1.61 (1.67) | -3.24 (0.76) | -0.61 (1.34) |
| **Key Stage 2** |  |  |  |  |  |
| ***English*** |  |  |  |  |  |
| Achieving expected level | 2008 (22.6%) | 1342 (29.7%) | 2319 (32.2%) | 33 (1.4%) | 10023 (56.4%) |
| Not achieving expected level | 1785 (20.1%) | 797 (17.7%) | 1449 (20.1%) | 51 (2.1%) | 4374 (24.6%) |
| Not assessed | 5080 (57.3%) | 2375 (52.6%) | 3436 (47.7%) | 2298 (96.5%) | 3360 (18.9%) |
| Standardised assessment score |  |  |  |  |  |
| Mean (SD) | -0.72 (1.18) | -0.46 (1.15) | -0.47 (1.14) | -1.27 (1.48) | -0.25 (2.09) |
| ***Maths*** |  |  |  |  |  |
| Achieving expected level | 2169 (24.4%) | 1234 (27.3%) | 2414 (33.5%) | 29 (1.2%) | 9775 (55.0%) |
| Not achieving expected level | 1667 (18.8%) | 889 (19.7%) | 1362 (18.9%) | 46 (1.9%) | 4609 (26.0%) |
| Not assessed | 5037 (56.8%) | 2391 (53.0%) | 3428 (47.6%) | 2307 (96.9%) | 3373 (19.0%) |
| Standardised assessment score |  |  |  |  |  |
| Mean (SD) | -0.70 (1.23) | -0.68 (1.18) | -0.50 (1.18) | -1.20 (1.50) | -0.37 (1.13) |

**Table S7.** Educational attainment outcomes across primary school assessments for children who sat all primary school assessments.

|  | **No ND before school**  **n (%)** | **ND before school**  **n (%)** | **Overall cohort**  **n (%)** |
| --- | --- | --- | --- |
| **Total** | 2119434 (100%) | 34462 (100%) | 2153896 (100%) |
| **Early Years Foundation Stage Profile** |  |  |  |
| ***Good Level of Development (GLD)*** |  |  |  |
| Achieving expected level | 1246730 (58.8%) | 14353 (41.6%) | 1261083 (58.5%) |
| Not achieving expected level | 872704 (41.2%) | 20109 (58.4%) | 892813 (41.5%) |
| ***English*** |  |  |  |
| Achieving expected level | 1497848 (70.7%) | 18640 (54.1%) | 1516488 (70.4%) |
| Not achieving expected level | 621586 (29.3%) | 15822 (45.9%) | 637408 (29.6%) |
| Standardised assessment score |  |  |  |
| Mean (SD) | 0.08 (0.91) | -0.32 (1.02) | 0.07 (0.85) |
| ***Maths*** |  |  |  |
| Achieving expected level | 1710325 (80.7%) | 23314 (67.7%) | 1733639 (80.5%) |
| Not achieving expected level | 409109 (19.3%) | 11148 (32.3%) | 420257 (19.5%) |
| Standardised assessment score |  |  |  |
| Mean (SD) | 0.08 (0.89) | -0.28 (1.03) | 0.07 (0.89) |
| **Key Stage 1** |  |  |  |
| ***English*** |  |  |  |
| Achieving expected level | 1469177 (69.3%) | 18288 (53.1%) | 1487465 (69.1%) |
| Not achieving expected level | 650257 (30.7%) | 16174 (46.9%) | 666431 (30.9%) |
| Standardised assessment score |  |  |  |
| Mean (SD) | 0.10 (0.85) | -0.27 (0.96) | 0.09 (0.85) |
| ***Maths*** |  |  |  |
| Achieving expected level | 1725416 (81.4%) | 22852 (66.3%) | 1748268 (81.2%) |
| Not achieving expected level | 394018 (18.6%) | 11610 (33.7%) | 405628 (18.8%) |
| Standardised assessment score |  |  |  |
| Mean (SD) | 0.09 (0.89) | -0.29 (1.01) | 0.09 (0.89) |
| **Key Stage 2** |  |  |  |
| ***English*** |  |  |  |
| Achieving expected level | 1667383 (78.7%) | 23014 (66.8%) | 1690397 (78.5%) |
| Not achieving expected level | 452051 (21.3%) | 11448 (33.2%) | 463499 (21.5%) |
| Standardised assessment score |  |  |  |
| Mean (SD) | 0.03 (0.98) | -0.34 (1.12) | 0.02 (0.98) |
| ***Maths*** |  |  |  |
| Achieving expected level | 1702542 (80.3%) | 23156 (67.2%) | 1725698 (80.1%) |
| Not achieving expected level | 416892 (19.7%) | 11306 (32.8%) | 428198 (19.9%) |
| Standardised assessment score |  |  |  |
| Mean (SD) | 0.01 (0.99) | -0.41 (1.15) | 0.01 (0.99) |

**Figure S3.** Proportion of children with subgroups of hospital-recorded neurodisability (ND) achieving nationally expected levels across English primary school assessments.

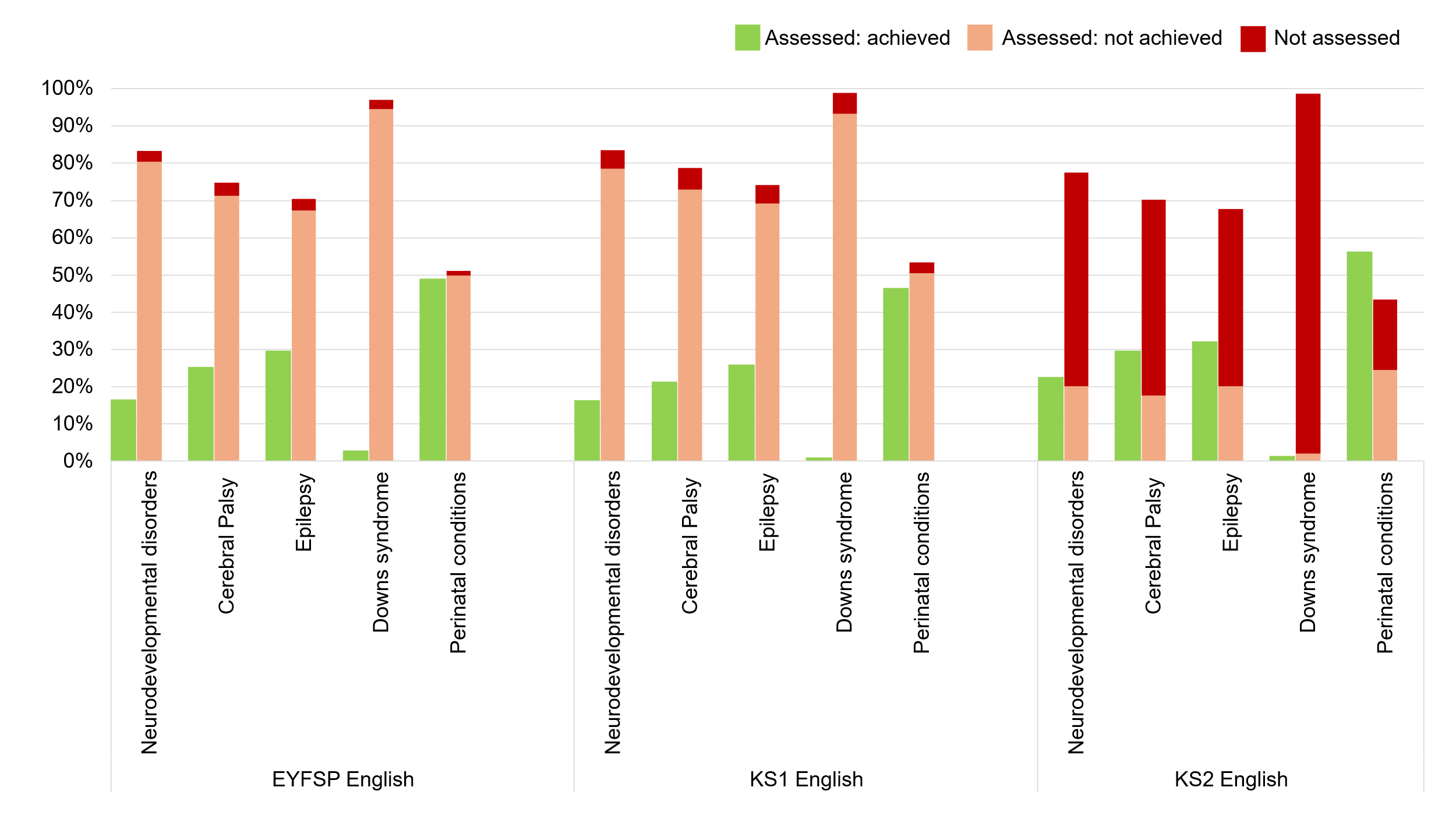

*EYFSP: Early Years Foundation Stage Profile. KS1 = Key Stage 1. KS2 = Key Stage 2. Subgroups of hospital-recorded neurodisability defined before Reception start (age 4/5)*
